# Analysis of East Asian Parkinson’s Disease Genomes Identifies Novel Susceptibility Loci and Functional Regulatory Variation

**DOI:** 10.64898/2026.09.03.26362212

**Authors:** Qiaoyang Sun, Ebonne Yulin Ng, Tzi Shin Toh, Kai Shi Lim, James Jia Dong Wang, Thomas Welton, Louis Chew Seng Tan, Kumar Manharlal Prakash, Ling-Ling Chan, Jia Nee Foo, Andrew Singleton, Laurel Screven, Hampton Leonard, Mike A. Nalls, Azlina Ahmad-Annuar, Yi Wen Tay, Han-Joon Kim, Jung Hwan Shin, Manabu Funayama, Nobutaka Hattori, Taku Hatano, Jee-Young Lee, Beomseok Jeon, Cheng-Hsuan Li, Sung-Pin Fan, Pin-Shiuan Chen, Chin-Hsien Lin, Shen-Yang Lim, Ai-Huey Tan, Eng-King Tan, The Global Parkinson’s Genetics Program (GP2)

## Abstract

**Background:** Parkinson disease (PD) is a genetically complex neurodegenerative disorder, but most genetic discoveries have been derived from populations of European ancestry, limiting the understanding of ancestry-specific genetic risk.

**Methods:** This GWAS included 5,825 East Asian participants (3,043 patients with PD and 2,782 controls). We then combined these results with data from two additional East Asian cohorts through meta-analysis, resulting in a total of 74,716 participants (15,603 patients with PD and 59,113 controls). To our knowledge, this represents the largest genetic study of PD in East Asian populations to date.

**Findings:** We identified two novel loci that were associated with PD in East Asian cohorts and reached genome-wide significance in the cross-ancestry meta-analysis (> 1.9 million subjects): TLE4 (lead variant rs10780320, P_meta_combined_ = 2.853×10^−^10) and HLA-V/HLA-G (lead variant rs11751333, P_meta_combined_ = 1.031×10-9). Integration of brain eQTL data identified an East Asian– specific intergenic variant at the LRRK2 locus (rs1388594) was significantly associated with LRRK2 expression in the basal ganglia. This association was replicated across 3 independent East Asian cohorts (P_gp2_EAS_= 8.71 × 10^−4^, P_sg_EAS_= 1.13 × 10^−5^, PTPMI_EAS = 4.86 × 10^−4^) and reached genome-wide significance in the East Asian meta-analysis (P_meta_EAS_ = 5.85 × 10^−10^; OR = 1.10, 95% CI: 1.07– 1.13). The variant was not associated with PD in populations of European ancestry (P = 0.08).

**Interpretation:** These findings improve our understanding of the genetics of PD across different ancestry groups and highlight the need to include people from different backgrounds in genetic studies to identify ancestry-specific risk variants.

## Introduction

Parkinson’s disease (PD) is a progressive neurodegenerative disorder marked by characteristic motor symptoms, including bradykinesia, rigidity, and resting tremor, together with a diverse range of non-motor manifestation^1^. As a common neurodegenerative disease worldwide, PD imposes a substantial and growing burden on aging populations, particularly in Asia, where demographic shifts are rapidly accelerating^2^. Genetic factors play a critical role in PD susceptibility. Over the past decade, genome-wide association studies (GWAS) have identified numerous risk loci, implicating key biological pathways such as lysosomal function, mitochondrial homeostasis, and immune regulation^3,4^. However, the majority of these discoveries have been derived from populations of European ancestry, raising concerns about their generalizability to other ethnic groups^4^. Differences in allele frequencies, linkage disequilibrium (LD) structure, and environmental exposures may contribute to population-specific genetic architectures and risk profiles^5^. East Asian (EAS) populations account for a substantial proportion of the global PD burden yet remain underrepresented in large-scale genetic studies. Although recent efforts, including international consortia and regional biobanks, have begun to address this gap^6,7^, the discovery of novel loci and refinement of known associations in EAS cohorts remain limited.

In this study, we performed a genome-wide association analysis in an East Asian cohort followed by a large-scale meta-analysis combining data from two additional major EAS cohorts. By integrating multiple cohorts with rigorous quality control and statistical approaches, we aim to identify novel PD-associated loci. Furthermore, we integrated the meta-analysis results with expression quantitative trait loci (eQTL) data to link genetic variants to gene expression and facilitate interpretation of their biological relevance. Our findings provide new insights into the genetic architecture of PD in EAS populations.

## Methods

### East Asian cohort in GP2 EAS

This study used genotype data from the Global Parkinson’s Genetics Program (GP2) Release 11 (https://gp2.org/), comprising 3,045 cases and 2,785 controls of East Asian ancestry. Genotype quality control was performed using PLINK v2.0. Variants with a call rate <95% and samples with genotype missingness >5% were excluded to ensure data integrity. Individual-level heterozygosity was assessed using the inbreeding coefficient (F statistic) in PLINK v2.0, and five samples with extreme heterozygosity (F < −0.15 or F > 0.15) were removed, resulting in a final dataset of 3,043 cases and 2,782 controls. At the variant level, Hardy–Weinberg equilibrium (HWE) filtering was applied, excluding variants with P < 1 × 10^−4^. Relatedness was evaluated using the KING algorithm, with a threshold of 0.0884 to remove pairs of individuals related up to the third degree. Ancestry inference was performed using GenoTools^8^, based on reference panels from the 1000 Genomes Project, the Human Genome Diversity Project, and the Ashkenazi Jewish dataset. The demographic and clinical characteristics are expressed as mean ± standard deviation (SD). A p-value <0.05 was regarded as statistically significant for non-GWAS comparisons. This study was conducted following the ethical standards of the relevant institutional and national research committees and was approved by the Operations and Compliance Working Group (OCWG) of GP2. Written informed consent was obtained from all participants at each participating site under locally approved ethical protocols.

### Genome-wide Association Analysis

Genome-wide association analysis was conducted using logistic regression implemented in PLINK2. Variants were restricted to those with MAF ≥ 0.01 and minor allele count (MAC) ≥ 20 prior to association testing. Association models were adjusted for sex and the top 10 principal components to control for population stratification. The firth-fallback option was applied to enable automatic use of Firth logistic regression when standard logistic regression failed to converge, particularly for low-frequency variants.

### East Asian and Cross Ancestry Meta-analysis

We first performed an East Asian GWAS meta-analysis by incorporating two additional large East Asian GWAS datasets: the Singapore (SG) GWAS dataset including 6,724 PD cases and 24,851 controls^9^, and the Taiwan Precision Medicine Initiative (TPMI) GWAS dataset comprising 5,836 PD cases and 31,480 controls^10^. For the SG GWAS dataset which is provided in the hg19 reference build, genomic coordinates were harmonized to hg38 using LiftOver. When multiple variants mapped to the same genomic position after conversion, only the variant with the smallest P value was retained to avoid redundancy and ensure a single representative signal per locus. Meta-analysis was conducted using METAL under a fixed-effects inverse-variance weighted model^11^. Only variants present in at least two datasets and variants exhibiting no substantial heterogeneity (I^2^ < 80%) were retained. Genomic inflation was assessed using the sample-size–adjusted inflation factor (λ_1000_). LD estimation was conducted using the EAS reference panel from the 1000 Genomes Project. Independent loci were defined using LD clumping with an r^2^ threshold of 0.6 within 250 kb windows. A locus was considered novel if it lay outside ±250 kb of previously reported regions^9,10,12-15^ and showed no evidence of LD with known variants. Genome-wide significant and suggestive loci were defined as variants with P < 5 × 10^−8^ and P < 5 × 10^−6^, respectively.

For further validation of genome-wide associated signals, we expanded the meta-analyses (total of > 1.9 million subjects) to include the GP2 European ancestry cohort comprising 63,555 cases, 17,700 proxy cases, and 1,746,386 controls.

### Functional Annotation and Enrichment Analysis

To gain biological insight into the identified association signals, we performed functional annotation and enrichment analyses using publicly available genomic resources. eQTL-based gene prioritization was conducted using data from the GTEx Project via FUMA^16^. The GTEx v8 dataset includes 13 brain tissues: amygdala, anterior cingulate cortex (BA24), caudate (basal ganglia), cerebellar hemisphere, cerebellum, cortex, frontal cortex (BA9), hippocampus, hypothalamus, nucleus accumbens (basal ganglia), putamen (basal ganglia), spinal cord (cervical c-1), and substantia nigra.

## Results

### GWAS analysis of GP2 EAS

After stringent quality control of variants and samples, 5,825 individuals from the EAS cohort of the GP2 dataset were retained, including 3,043 PD cases and 2,782 controls cases (mean [SD] age: 67.8 ± 9.9 years; age at onset: 59.2 ± 11.5 years; 52.6% men; 7.1% with a family history) and 2,782 controls (mean [SD] age: 62.5 ± 11.5 years; 67.5% men) (Table. 1). Genome-wide association analyses were conducted using logistic regression, adjusting for sex and the first ten principal components (PC1–PC10) to account for population stratification. The sample size–adjusted genomic inflation factor was low (λ_1000_ = 1.038), and the quantile–quantile (QQ) plot showed no evidence of substantial inflation (Supplementary Fig. 1).

**Table 1.** Demographic Characteristics of Patients With Parkinson Disease and Control Participants.

| phenotype | Number | Age_sampling | Age_onset | Male | Female | Family_history |
| --- | --- | --- | --- | --- | --- | --- |
| Control | 2782 | 62.5 ± 11.5 | NaN ± NA | 1878<br>(67.5%) | 904 (32.5%) | 11 (0.4%) |
| PD | 3043 | 67.8 ± 9.9 | 59.2 ± 11.5 | 1601<br>(52.6%) | 1442<br>(47.4%) | 216 (7.1%) |

Following LD-based clumping of association signals, we identified 10 independent genome-wide significant loci (P < 5 × 10^−8^), all of which have been previously reported. Of these, nine loci were located on chromosome 4, with the strongest signal represented by the lead SNP rs6830166 near *SNCA* (P = 2.8 × 10^−20^). One additional locus was identified on chromosome 1, with lead SNP rs1775146 near *RAB29 and PM20D1* genes. Each locus exhibited clusters of correlated variants, supporting robust and well-defined association signals (Supplementary Table 1).

In addition to genome-wide significant loci, 27 independent loci showed suggestive evidence of association (P < 5 × 10^−6^). Among these, 12 were previously reported, while 15 appear to be novel. These loci were distributed across multiple chromosomes, including chromosomes 4, 5, 7, 9, 10, 12, 15, 16, and 19. Most loci were supported by multiple variants across significance thresholds, whereas others were driven by a limited number of SNPs—such as loci on chromosomes 3, 7, 15, and 19—suggesting more isolated association signals (Fig. 2A; Supplementary Table 1).

**Figure 1.**
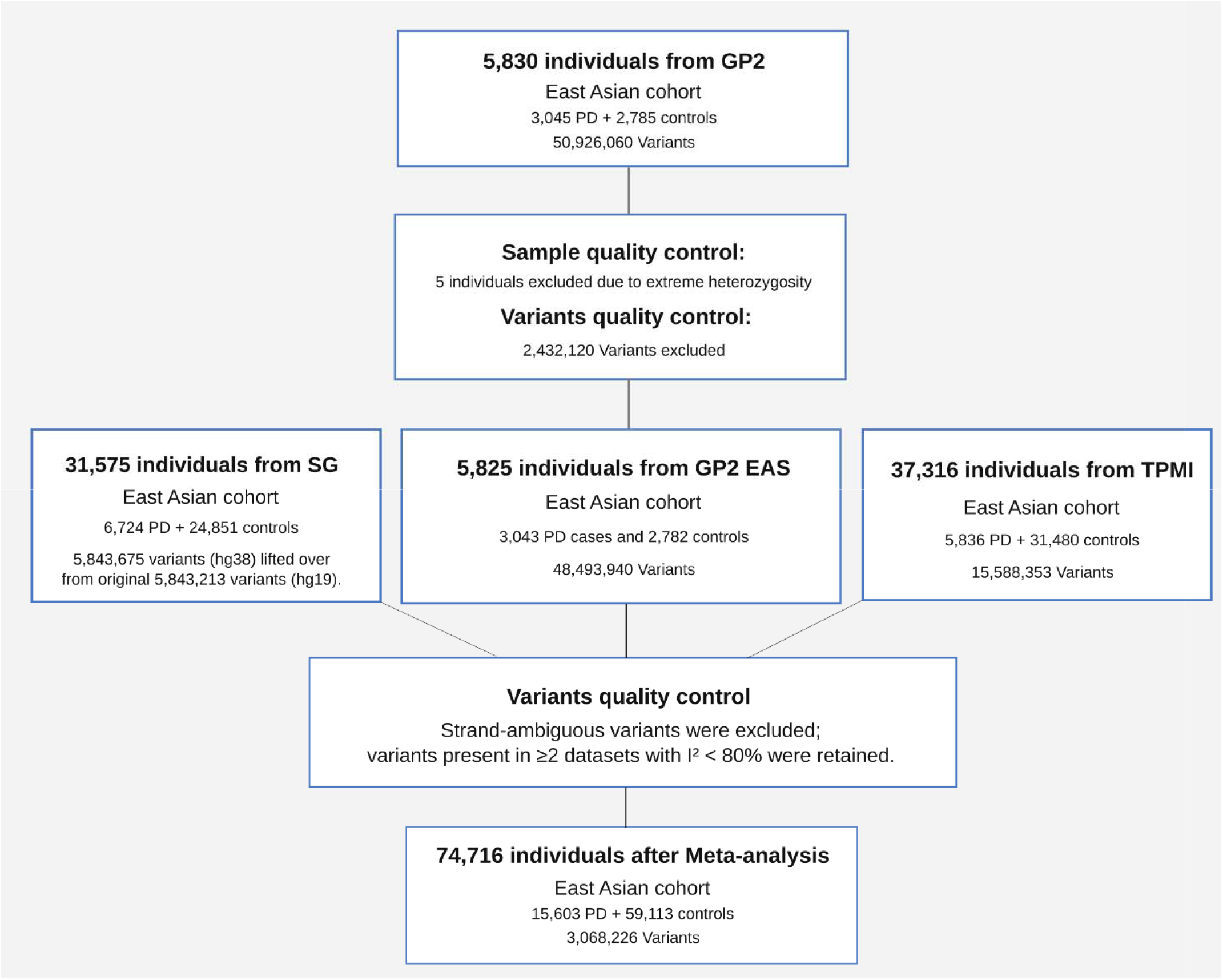
Study workflow. Schematic overview of the study design.

**Figure 2.**
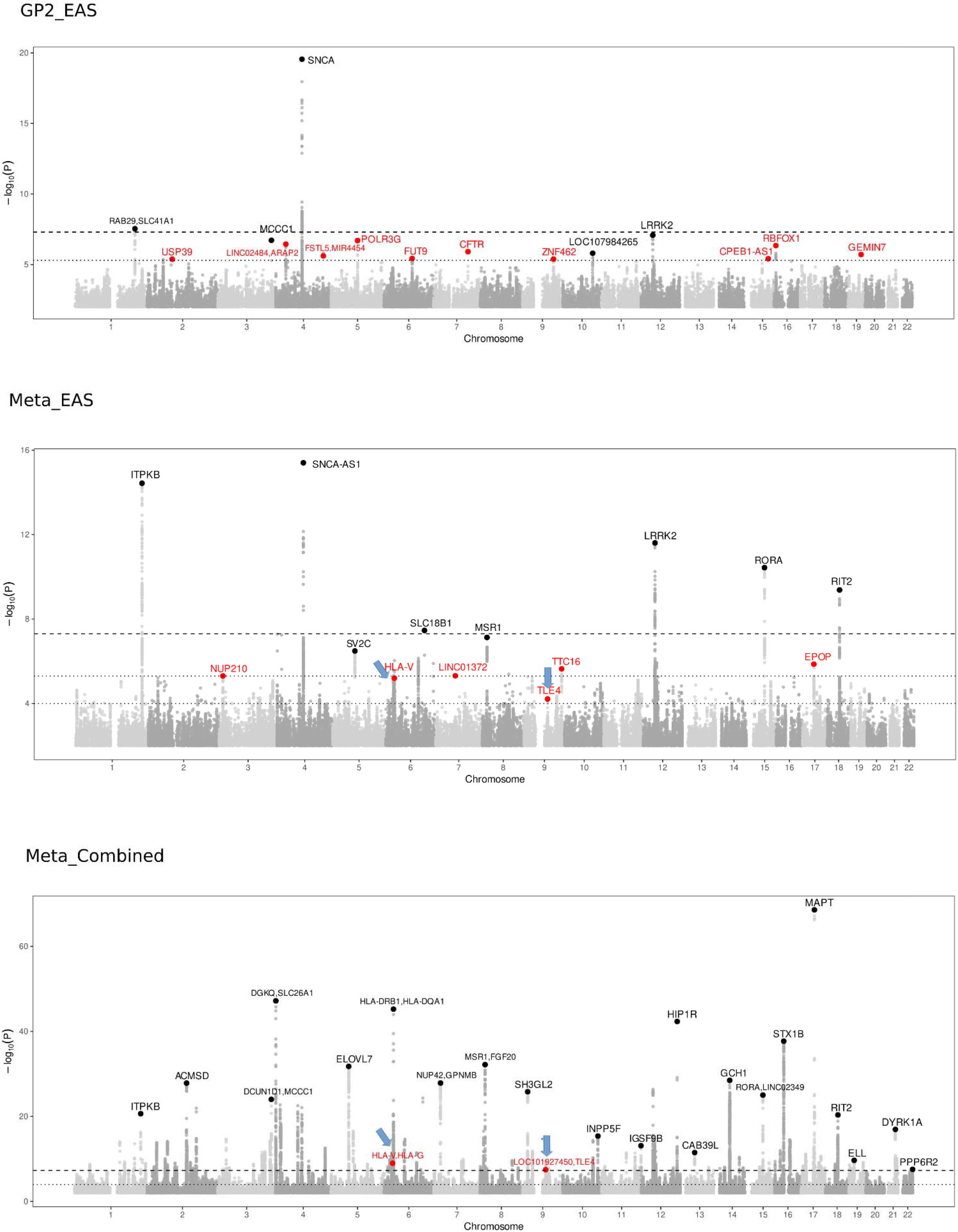
Manhattan plots of association signals across analyses. Genome-wide significance (P < 5 × 10^−8^) and suggestive significance (P < 5 × 10^−6^) thresholds are indicated. Lead SNPs at novel loci are highlighted in red, whereas those at previously reported loci are shown in black. An additional threshold line (P < 1 × 10^−4^) is included in the Meta_EAS plot to highlight loci showing suggestive evidence of association. Blue arrows indicate the two novel loci that were suggestively associated in the Meta_EAS analysis and reached genome-wide significance in the Meta_combined analysis.

### Meta-Analysis of EAS GWAS (Meta_EAS)

To increase power for locus discovery, we performed a meta-analysis combining our dataset with two additional large EAS GWAS datasets, yielding a total of 15,603 cases and 59,113 controls (hereafter referred to as Meta_EAS). Following meta-analysis, genomic inflation was minimal (λ_1000_ = 1.003), and the QQ plot likewise indicated well-controlled population structure (Supplementary Fig. 1). A total of 25 loci reached genome-wide significance (P < 5 × 10^−8^), while 35 loci showed suggestive evidence of association (P < 5 × 10^−6^) (Fig. 2; Supplementary Table 2). All 25 genome-wide significant loci mapped to previously reported regions, including established signals at *ITPKB* (rs12095028), *SNCA-AS1* (rs1023777), *LRRK2* (rs10506151), *SLC18B1* (rs41286192), and *RORA* (rs2243453). Among the 35 suggestive loci, 28 overlapped with previously reported regions, including *SV2C* (rs246814) and *MSR1* (rs635052). The remaining seven loci were considered potentially novel (Fig. 2), including regions near *NUP210* (rs3732671), *LINC01372* (rs10280589), *TTC16* (rs3802357), *HLA-E* (rs371172638), *PACRG (rs9458749), PIP4P2* (rs7016498), and *EPOP* (rs720528).

### Validation of the novel loci by further combining a Cross-Ancestry GWAS dataset (Meta_combined)

To further validate these novel association signals, we performed a cross-ancestry meta-analysis by combining the EAS meta-analysis (Meta_EAS) with the GP2 European GWAS dataset (63,555 cases, 17,700 proxy cases, and 1,746,386 controls), hereafter referred to as Meta_combined. Only variants present in both datasets and showing no substantial heterogeneity (I^2^ < 80%) were retained. (I^2^ < 80%) were retained. The expanded meta-analysis identified additional association signals (Supplementary Table 3). We then applied the following criteria to identify loci that showed nominal evidence of association in the EAS-specific meta-analysis and reached genome-wide significance in the combined analysis: (1) the association signal in the EAS cohort reached a suggestive significance threshold (P < 1 × 10^−4^); (2) the lead SNPs from the EAS-specific and combined meta-analyses were located within the same LD block (r^2^ > 0.6); and the lead SNPs were within 250 kb of each other. Applying these criteria, we identified two independent novel loci (Fig. 3; Supplementary Tables 4 and 5). The first locus was located in the *TLE4* (Transducin-Like Enhancer of Split 4) region. The lead variant, rs10780320 (chr9:79,711,197), reached genome-wide significance in the combined meta-analysis (P _meta_combined_= 2.853 × 10^−10^) (Fig. 3). In the EAS-specific meta-analysis, this locus showed suggestive evidence of association, with rs4877149 (chr9:79,728,887) identified as the lead variant (P _meta_EAS_= 6.118 × 10^−5^).

**Figure 3.**
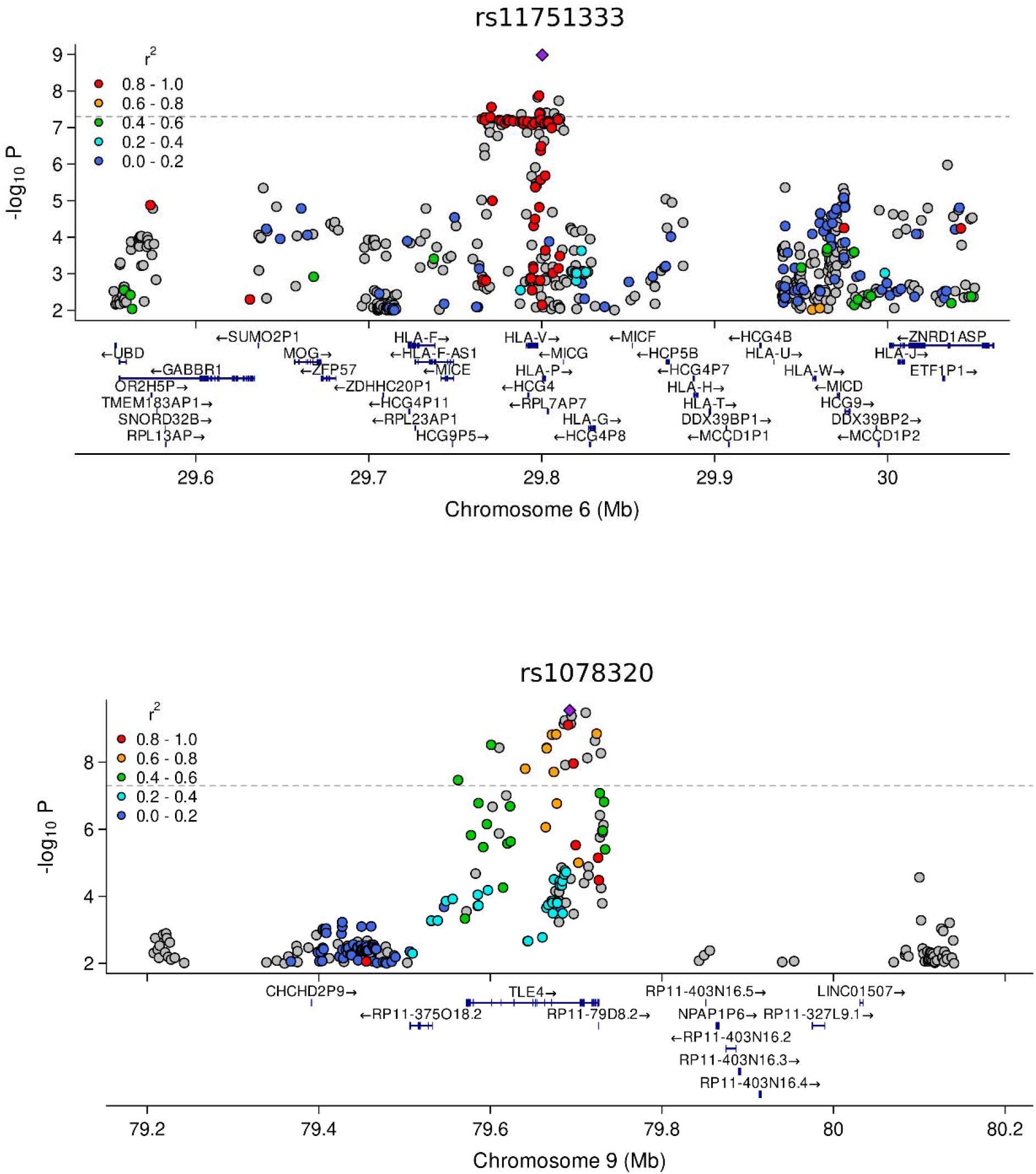
Novel PD risk loci identified in the Meta_combined dataset. Regional association plots of the two novel loci (*TLE4* and *HLA*) are shown. Both loci exhibited suggestive evidence of association in the East Asian meta-analysis (Meta_EAS) and reached genome-wide significance in the combined meta-analysis (Meta_combined). The lead SNP and surrounding variants are displayed, with variants colored according to their linkage disequilibrium (LD; r^2^) with the lead SNP. LD estimates were calculated using the East Asian reference panel from the 1000 Genomes Project.

The second novel locus was identified in the *HLA-V/HLA-G* region. The lead variant rs11751333 (chr6:29,800,360) reached genome-wide significance in the combined meta-analysis (P _meta_combined_= 1.031 × 10^−9^). In the EAS-specific meta-analysis, rs72492267 (chr6:29,809,849) was the lead variant at this locus and showed suggestive evidence of association (P _meta_EAS_ = 6.346 × 10^−6^) (Fig. 3B,C).

Although the lead SNPs differed between the EAS-specific and combined meta-analyses, the variants identified in each analysis showed evidence of association in the other. Specifically, the EAS-specific lead variants reached genome-wide significance in the combined meta-analysis (rs72492267 at the *HLA locus*, P _meta_combined_= 1.84 × 10^−8^; rs4877149 at the *TLE4 locus*, P _meta_combined_= 5.40 × 10^−10^). Similarily, the combined meta-analysis lead variants remained significant associated in the EAS-specific analysis (rs11751333 at the *HLA locus*, P _meta_EAS_= 8.20 × 10^−5^; rs10780320 at the *TLE4 locus*, P _meta_EAS_ = 3.94 × 10^−4^) (Supplementary Table 6). The differences in lead SNPs between Meta_EAS and Meta_combined datasets likely reflect differences in LD patterns between East Asian and European populations. However, the paired lead SNPs are located close to each other within the same LD blocks, suggesting that they represent the same underlying association signals. Overall, these findings support the associations identified in the EAS-specific analysis, with additional samples in the combined meta-analysis further increasing statistical confidence.

### Prioritization of Intergenic Variants Using eQTL Integration

We next prioritized variants from the Meta_EAS analysis using the FUMA platform based on their regulatory effects on gene expression. Tissue expression analysis, with GTEx data across 53 tissue types, showed the strongest enrichment in brain tissues (Fig. 4A). Gene-based testing with MAGMA identified three genes with the most significance—*ITPKB, SNCA*, and *LRRK2* (Fig. 4B). Consistent with this, eQTL analysis showed detectable expression signals at these loci. Specifically, multiple SNPs within the locus on chromosome 4 (4:90,453,241–91,113,666) were associated with *MMRN1* expression, while the locus on chromosome 1 was linked to *ADCK3* expression (Supplementary Fig. 2). In contrast, the *LRRK2* locus showed a single clear variant with a significant effect on gene expression. Although multiple variants were present in this region, only the intronic variant rs1388594 (Fig. 4C) was significantly associated with LRRK2 expression, specifically in the caudate nucleus. This highlights rs1388594 as the likely regulatory driver at this locus in the East Asian population. Importantly, rs1388594 was consistently replicated across all three East Asian cohorts included in our meta-analysis—GP2_EAS GWAS (P = 8.71 × 10^−4^, OR = 1.15, 95% CI: 1.06–1.26), SG_EAS GWAS (P = 1.13 × 10^−5^, OR = 1.11, 95% CI: 1.06–1.16), and TPMI_EAS GWAS (P = 4.86 × 10^−4^, OR = 1.07, 95% CI: 1.03–1.12) —and reached genome-wide significance in the combined analysis (P = 5.85 × 10^−10^, OR = 1.10, 95% CI: 1.07–1.13). In contrast, no significant association of rs1388594 with PD was observed in European-ancestry samples within GP2 (P = 0.084), suggesting a potential East Asian–specific risk signal. Notably, rs1388594 is independent of previously reported *LRRK2* variants, including Asian risk variants G2385R (rs34778348) and R1628P (rs33949390) (Supplementary Table 7).

**Figure 4.**
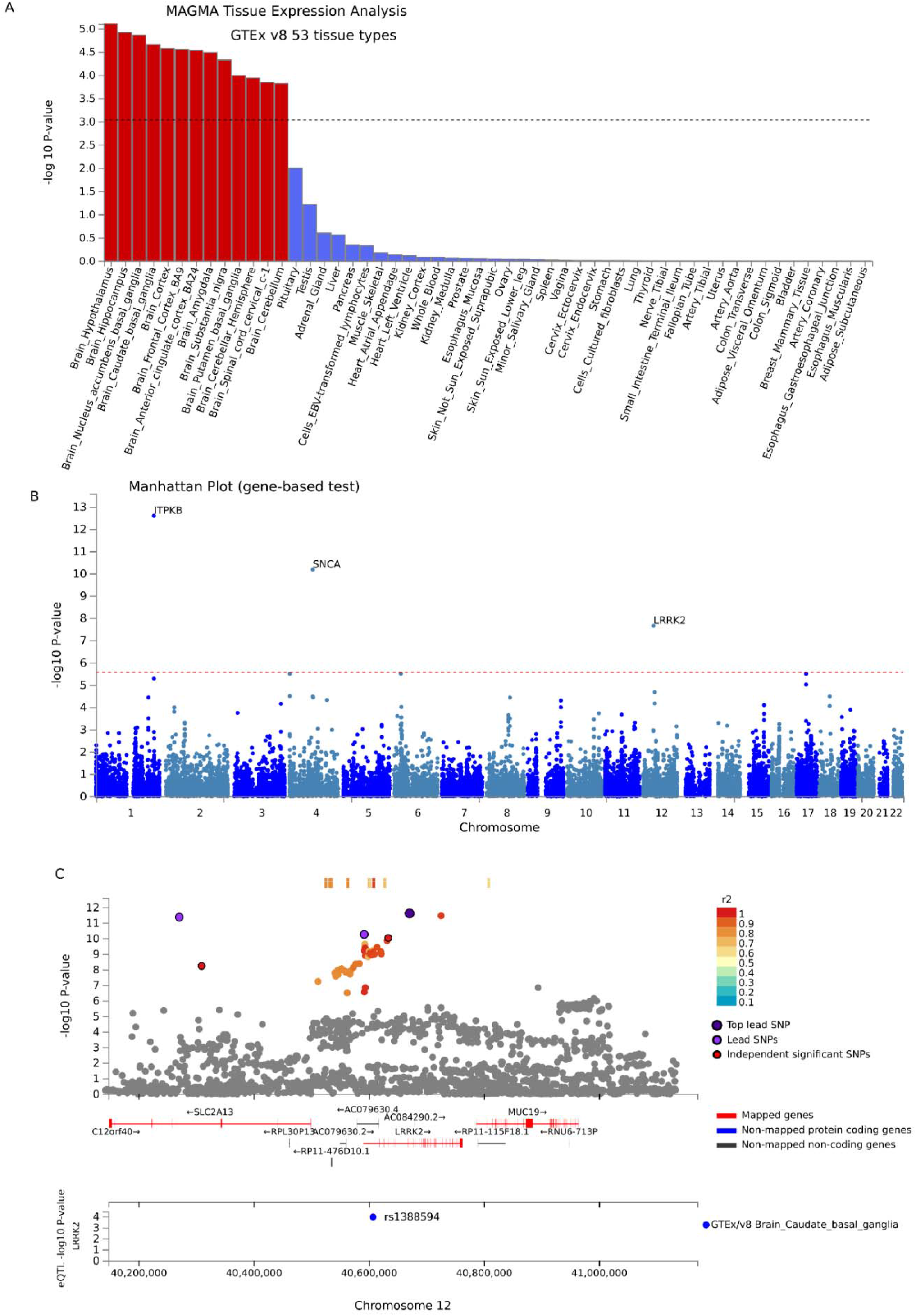
eQTL analysis identifying rs1388594 as the single variants within the loci in Meta_EAS which influencing *LRRK2* expression in caudate nucleus. (A) MAGMA analysis to identify tissue-specific enrichment of gene-level associations. (B) Gene-based Manhattan plot from MAGMA. A total of 18,416 protein-coding genes were tested, with genome-wide significance set at *P* = 2.715 × 10^−6^ (red dashed line). (C) Regional plot showing GWAS and eQTL signals across 13 brain tissues from the GTEx Project. The intronic variant rs1388594 was the only variant significantly associated with LRRK2 expression, with the association observed specifically in the caudate nucleus. Significant eQTLs are displayed, with filtered variants shown in grey. Gene colors indicate annotation categories: red = mapped genes; blue = non-mapped protein-coding genes; dark grey = non-coding genes.

## Discussion

PD is a complex neurodegenerative disorder in which genetic factors play a central role in disease susceptibility and pathogenesis^1^. Although numerous risk loci have been identified, with most discoveries derived from populations of European ancestry^17^. In contrast, the genetic architecture of PD remains incompletely understood in underrepresented populations, including East Asians. This knowledge gap is partly due to the limited availability of GWAS data from East Asian populations, which reduces the statistical power to detect disease-associated variants. Meta-analysis provides an effective strategy to overcome this limitation by combining data from multiple independent studies, thereby increasing statistical power and improving the detection of genetic association signals. By integrating evidence across cohorts, meta-analysis enhances the robustness, reliability, and precision of genetic findings, facilitating the discovery and validation of PD risk variants.

In this study, we performed a GWAS in an East Asian cohort and conducted a meta-analysis incorporating two additional large East Asian PD cohorts, resulting in a total sample size of 74,716 individuals, the largest EAS PD meta-analysis to date^9,10^. We replicated 53 previously reported loci associated with PD. In addition, we identified two novel loci, including one on chromosome 9 and one on chromosome 6, that showed suggestive evidence of association in the EAS-specific analysis and reached genome-wide significance in the combined meta-analysis. The chromosome 9 locus mapped to *TLE4*, which encodes a transcriptional corepressor expressed in both developing and mature cortical neurons. It is required for the embryonic acquisition and postnatal maintenance of corticothalamic projection neuron identity. Loss of *TLE4* function disrupts this identity, leading these neurons to acquire characteristics of alternative neuronal subtypes, such as subcerebral projection neurons^18^. Given its role in neuronal differentiation and transcriptional regulation, genetic variation at this locus may implicate cortical regulatory mechanisms in the pathogenesis of PD, although further functional validation is required. The locus on chromosome 6 reside near the major histocompatibility complex (MHC), a highly polymorphic and gene-dense region characterized by extensive linkage disequilibrium^19,20^. This complex LD structure may obscure causal variants and complicate interpretation of independent signals, so these findings should be interpreted cautiously. Given the increasing evidence for immune involvement in PD^21,22^, replication in independent Asian cohorts will be important to confirm the robustness and functional relevance of these associations.

Another key strength of this study is the identification of regulatory variants that are unique to or enriched in East Asian populations, providing an opportunity to uncover disease mechanisms that may not be captured in studies of European populations alone. comparing these ancestry-specific regulatory variants with risk variants identified in European cohorts, we can determine whether distinct genetic signals converge on shared genes, regulatory elements, or biological pathways. By integrating GWAS results with eQTL data from 13 human brain regions, we identified putative functional variants and discovered rs1388594 as a novel East Asian–specific regulatory signal at the LRRK2 locus. Importantly, rs1388594 is independent of previously reported LRRK2 risk variants, including the Asian-specific variants G2385R (rs34778348) and R1628P (rs33949390). *LRRK2* is one of the most consistently replicated genetic factors for PD^23-26^ and encodes a multifunctional kinase involved in vesicle trafficking, autophagy, endolysosomal dynamics, and cytoskeletal regulation, all of which are essential for neuronal homeostasis ^27-29^. While coding pathogenic mutations such as G2019S and R1441G are known to increase kinase activity^30^, emerging evidence suggests that Asian-prevalent risk variants such as G2385R also increase kinase activity, albeit with a smaller effect size^31^. However, the contribution of non-coding regulatory variation to *LRRK2* expression remains less well characterized, despite being highlighted for some years^32^. Importantly, the identified variant showed consistent direction and magnitude of effect across all three East Asian GWAS datasets, but did not demonstrate a significant association in European ancestry datasets. This pattern suggests a potential East Asian–specific regulatory effect at the *LRRK2* locus, highlighting ancestry-dependent genetic architecture in PD.

In conclusion, our findings expand the genetic architecture of PD in East Asian populations by identifying novel and ancestry-specific risk loci, underscoring the importance of diverse populations in uncovering disease biology.

## Data Availability

The data generated in this study have been deposited in GP2 and will be made available through the GP2 platform (https://gp2.org) in accordance with GP2 data-sharing policies.

## Declaration of generative AI and AI-assisted technologies in the writing process

During the preparation of this work, the authors used ChatGPT to check the language. After using this tool/service, the authors reviewed and edited the content as needed, taking full responsibility for the content of the publication.

## Acknowledgements

This research is supported by the Singapore Ministry of Health’s National Medical Research Council under its < Open Fund Large Collaborative Grant (MOH-OFLCG24may-0004) and Singapore Translational Research (STaR) Investigator Award (to EK-T). This project is also supported by the Global Parkinson’s Genetics Program (GP2; https://gp2.org). GP2 is funded by the Aligning Science Across Parkinson’s (ASAP) initiative and implemented by The Michael J. Fox Foundation for Parkinson’s Research (MJFF). For a complete list of GP2 members see https://doi.org/10.5281/zenodo.7904831.

## Data Sharing Statement

### Data Availability

Data used in the preparation of this article were obtained from the Global Parkinson’s Genetics Program (GP2; https://gp2.org). Specifically, we used Tier 1 and Tier 2 data from GP2 Release 11. The data generated in this study have been deposited in GP2 and will be made available through the GP2 platform (https://gp2.org) in accordance with GP2 data-sharing policies.

### Code Availability

All code generated for this article, and the identifiers for all software programs and packages used, are available on GitHub [https://github.com/GP2code/GP2-EAS-GWAS] and Zenodo DOI: 10.5281/zenodo.20348849.

## Contributors

EKT, CHL, SYL, and AHT conceptualised the study. EYN, TST, KSL, JJW, TW, LCST, KMP, LLC, JNF, AS, LS, HL, MAN, AA-A, YWT, HJK, JHS, MF, NH, TH, JYL, BJ, CHL, SPF, and PPC contributed to sample collection, data curation, analysis, and interpretation. QS drafted the manuscript. All authors reviewed and revised the manuscript and approved the final version.

